# SACCHARIN TRANSIT TIME IN EXCLUSIVE E-CIGARETTES AND HEATED TOBACCO PRODUCTS USERS: A CROSS-SECTIONAL STUDY

**DOI:** 10.1101/2020.10.21.20216630

**Authors:** R. Polosa, R. Emma, F. Cibella, M. Caruso, G. Conte, F. Benfatto, S. Ferlito, A. Gulino, M. Malerba, P. Caponnetto

## Abstract

Tobacco smoking impairs mucociliary clearance (MCC) efficiency as shown by prolonged saccharin test transit time (STTT). Avoiding exposure to tobacco smoke from combustible cigarettes may restore MCC function and former smokers have been shown to exhibit similar STTT as never smokers. The impact of switching from smoking to combustion-free tobacco products such as e-cigarettes (ECs) and heated tobacco products (HTPs) on STTT is not known. We report STTT of exclusive EC and HTP users. Test results were compared to those obtained in current, former, and never smokers.

STTT were obtained from 39 current, 40 former, 40 never smokers and from 20 EC and 20 HTP users. Comparison of STTT values showed significant difference among the five study groups (p< 0.00001) with current smokers having a median (IQR) STTT of 13.15 min, which was significantly longer compared to that of all other study groups. In particular, compared to former (7.26 min) and never smokers (7.24 min), exclusive EC users and exclusive HTP users had similar STTT at 7.00 and 8.00 min respectively.

Ex-smokers who have switched to exclusive regular use of combustion-free nicotine delivery systems (i.e. ECs and HTPs) exhibit similar saccharin transit time as never and former smokers. This suggests that combustion-free nicotine delivery technologies are unlikely to have detrimental effects on MCC function.

## INTRODUCTION

In addition to causing cardiovascular diseases, cancer and chronic obstructive pulmonary diseases (COPD) (1,2), chronic exposure to a range of toxic chemicals in the cigarette smoke can cause progressive structural damage and functional alterations of the airways, with loss of cilia (3,4), reduced ciliary beating (5,6), and airway epithelial mucus cell hyperplasia causing mucus hypersecretion (7,8). Tobacco combustion in conventional cigarettes is known to release a multitude of harmful and potentially harmful chemical constituents including phenol, formaldehyde, and acrolein (9), which have been shown to be cilia-toxic (10). Disruption of the mucociliary clearance function may contribute to inflammation and obstruction of the small airways (11), and increased susceptibility to respiratory infections (12-14).

Abstaining from tobacco smoking may reduce structural damage and restore cilia-mucus interaction. Former smokers have been shown to exhibit similar MCCTT as never smokers (15) and smoking cessation studies have demonstrated that MCC impairment can be rapidly reversed in quitters (16,17). The demonstration that the cilia-mucus functional framework of smokers can be restored soon after stopping exposure to smoke toxicants suggests that measurement of MCCTT can be used as a sensitive biomarker of physiological effect for the detection of early respiratory health changes in smoking cessation studies and switching trials of combustion-free nicotine delivery systems (e.g. e-cigarettes, heated tobacco products).

Products that do not require combustion to deliver nicotine such as e-cigarettes (ECs) and heated tobacco products (HTPs) are substituting conventional cigarettes globally (18). Compared to conventional cigarettes, they offer substantial reduction in exposure to harmful and potentially harmful chemical constituents including phenol, formaldehyde, and acrolein (19-22) and for this reason combustion-free nicotine delivery systems are have been considered for smoking harm reduction (23).

Given that a significant reduction in combustion toxicants when stopping smoking reverses the impairment of the cilia-mucus functional structure, it is hypothesized that faster MCCTT can be shown in exclusive users of combustion-free nicotine delivery systems compared to current smokers. To test this hypothesis, we carried out Saccharin tests - a non-invasive, well tolerated and simply to perform method that measures MCCTT (24) - in a group of exclusive e-cigarettes and heated tobacco products users. Test results were compared to those obtained in current, former, and never smokers.

## METHODS

### Study Population

Adult smokers who attended a smoking cessation clinic (CPCT, Centro per la Prevenzione e Cura del Tabagismo of the University of Catania), or subjects contacted among hospital staff, or through social media were recruited to establish five separate study groups.

Study group 1 consisted of current smokers smoking ≥ 10 cigarettes per day, with an exhaled carbon monoxide (eCO) level of ≥ 7 ppm.

Study group 2 consisted of former smokers, who stopped smoking for at least 6 months after completing their smoking cessation program at CPCT in the previous 2 yrs, and were still abstinent with an eCO level of < 7 ppm.

Study group 3 consisted of never smokers, who reported having smoked less than 100 cigarettes in their lifetime (25) and with an eCO level < 7 ppm to exclude subjects significantly exposed to environmental cigarette smoke.

Study group 4 consisted of exclusive regular e-cigarette users, who completely stopped smoking for at least 3 months after switching to their e-cigarette and were still abstinent with an eCO level of < 7 ppm.

Study group 5 consisted of exclusive regular heated tobacco product users, who completely stopped smoking for at least 3 months after switching to their heated tobacco product and were still abstinent with an eCO level of < 7 ppm.

Study participants had to satisfy the following exclusion criteria:

- Any conditions that could impair cilia-mucus interaction or interfere with MCCTT measurements, such as:
  ▪ Recent (less than 14 days) hx of viral infection of the upper respiratory tract
  ▪ Conditions that may damage nasal mucosa (e.g. chronic rhinosinusitis, infectious rhinitis, allergic rhinitis, atrophic rhinitis, vasomotor rhinitis)
  ▪ Respiratory conditions that may interfere with MCCTT measurements (e.g. COPD, asthma, bronchiectasis, cystic fibrosis)
  ▪ Significant exposure to passive smoking (excludes current smokers)
  ▪ Significant environmental/occupational exposure to pollution or chemicals (e.g. living in proximity of areas characterized by heavy vehicles traffic, or by presence of industrial fumes; employment in chemical/metallurgy industries)
  ▪ Medications such as pain killers, sleeping pills, antihistamines
- Poor individual ability to detect sweetness (i.e. being below the 25 mm mark on the 0-100 mm VAS for sweetness intensity rating)
- Current use of e-cigarettes or heated tobacco products (for current, former and never smokers only)
- Pregnancy

The study was approved by the local Ethical Review Board (number 125/2019/empo, Comitato Etico Catania 1. AOU Policlinico Vittorio Emanuele) and participants gave written informed consent prior to participation in the study.

### Study Design

This is a cross-sectional study designed to assess MCCTT among five study populations: 1) current smokers; 2) former smokers; 3) never smokers; 4) exclusive EC users (ex-smokers); and 5) exclusive HTP users (ex-smokers). After a screening visit, subjects were invited to attend for a saccharin test.

At screening, eligibility criteria (socio-demographic data, medical history, medication usage, and tobacco products history) were verified. Potential participants were tested for exhaled CO and their ability to detect sweetness. Perception of sweetness intensity was rated by using a 0-100 mm VAS. After rinsing the mouth with tap water and wiping the tongue dry with a paper towel, subjects were instructed to smear a saccharine tablet (Mini-sweeteners; Hermesetas; Switzerland) all around the surface of their tongue. They then were asked to rate the intensity of sweetness perception on a 0-100 mm VAS. Sweetness intensity ratings ranged from ‘not at all sweet’ (at 0 mm) to ‘*extremely sweet*’ (at 100 mm). Anybody below the 25 mm mark on the VAS was excluded from participation. Eligible subjects were then invited to attend the Saccharin test visit. They were asked to refrain from drinking coffee/caffeinated drinks for at least 4 h prior to the study visit. Smokers were asked not to smoke, EC users not to vape, and HTP users not to puff their device for at least 1 h prior to the Saccharin test visit. At Saccharin test visits, eligibility criteria were verified once again. Before commencing the saccharin test, subjects’ nose was rinsed with warm saline (NaCl 0,9% solution). After asking participants to acclimatize at controlled environmental conditions (temperature 21-24°C; relative humidity 30-50%) for at least 45 min, Saccharin test transit times were measured.

### Saccharin Test Method

After nasal washing with warm saline, participants were invited to acclimatize in an examination room optimized for ambient temperature and humidity (i.e. temperature 21-24°C; relative humidity 40-60%). After 45 min acclimatization, participants were invited to slightly raise and tilt the head backwards. Whilst illumining a nostril (that indicated by the subject as the one allowing better nasal breathing – the same nostril will be used for all tests providing patency is maintained throughout study visits) with the medical headlight and widening it by using a nasal speculum, the research investigator (or ENT research nurse) identified the small crest that marks the tip of the inferior turbinate. The nipper clasping a saccharin tablet was guided through the speculum and the tablet was gently placed horizontally on the medial face of the inferior turbinate, about 1 cm behind its anterior end. The nipper and nasal speculum were withdrawn paying attention not to trigger any sneezing. Subjects were then invited to return their heads to a straight position and a chronometer was started. Subjects were asked to swallow some saliva a few times every minute until perceiving the “sweet taste” of saccharin. Subjects were instructed to avoid to sniff, sneeze, eat, drink, walk, talk, cough, scratch or blow their nose.

### Statistical Analysis

Based on data from previous Saccharin test studies comparing current, former and never smokers, we have estimated that a sample of at least 20 subjects for each group was adequate to obtain a power greater than 90% with a type-I error (alpha) smaller than 0.05 (5%) in an equivalence comparison. The Lower Limit of Normality (LLN) was calculated by computing the value corresponding to the mean + SD*1.64 from the distribution curve of the results of the MCCTT measurements in never smokers. Kolmogorov-Smirnov test was performed to assess the data distribution. Categorical data were summarized by counts and percentages; continuously distributed data, with symmetrical distribution, were summarized using the mean (standard error; SE); continuously distributed data, with skewed distribution, were summarized using the median (inter-quartile range; IQR). Study groups comparisons were carried out by Chi-square test, ANOVA and Kruskall-Wallis test for categorical, continuously symmetric and continuously skewed datasets, respectively. Moreover, cross-comparison between groups were calculated using pairwise Wilcoxon Test with Holm correction for multiple testing. All analyses were considered significant with a P-value< 0.05. R version 3.4.3 (2017-11-30) was utilized for data analysis and generation of graphs.

## RESULTS

### Study Participants

Complete analysis on the saccharin test was carried out in 159 subjects: 79F/80M with a median (IQR) age of 32 (25-42) years (**Table 1**). No significant differences were observed among the study groups, with the exception of exhaled eCO levels, which were significantly (p<0.0001) elevated in current smokers compared to never smokers, former smokers, exclusive HTP users and exclusive EC users.

**Table 1.** Demographic and Clinical Characteristics of Study Groups

|  | Never<br>Smokers | Current<br>Smokers | Former<br>Smokers | Exclusive<br>HTP users | Exclusive<br>ECIG users | *p value* |
| --- | --- | --- | --- | --- | --- | --- |
| **Age** | 32.5 (25-41) | 31 (24.5-44) | 33 (25.75-41.25) | 34.5 (26.5-44.75) | 33.5 (25.75-42) | 0.979 |
| **F/M** | 20/20 | 19/20 | 20/20 | 10/10 | 10/10 | 0.991 |
| **BMI** | 23.37 ± 3.23 | 25.07 ± 4.97 | 23.94 ± 4.39 | 25.22 ± 4.46 | 24.03 ± 4.76 | 0.092 |
| **Exhaled CO** | 3 (2-4) | 19 (15-22.5) | 2.5 (1.75-5.00) | 3 (2-5) | 3 (1.75-4.25) | <0.0001 |
| **Pack/Years** | NA | 13.5 (6.15-20.35) | 15 (5.06-25.35) | 16.25 (9.15-26.55) | 12 (6.95-27.95) | 0.954 |
| **Cigarettes/day** | NA | 15 (11-20) | 20 (15-25)* | 20 (12-25)* | 16 (11-20)* | 0.268 |
| **FTND** | NA | 6 (5-7) | NA | NA | NA | - |
| *Data are reported as mean ± SD, median (IQR), n/N unless otherwise stated.*<br>*BMI: body mass index; CO: carbon monoxide; FTND: Fagerstrom Test for Nicotine Dependence.*<br>*\*refers to previous smoking (prior to quitting)* |  |  |  |  |  |  |

### MCCTT Comparison between study groups

Comparison of MCCTT values showed significant difference among the five study groups (p< 0.00001); current smokers had a median (IQR) MCCTT of 13.15 (9.89-16.08) min, which was significantly longer compared to that of never smokers at 7.24 (5.73-8.73) min, former smokers at 7.26 (6.18-9.17) min, exclusive EC users at 7.00 (6.38-9.00) min, and exclusive HTP users at 8.00 (6.00-8.00) min (**Figure 1**).

**Figure 1.**
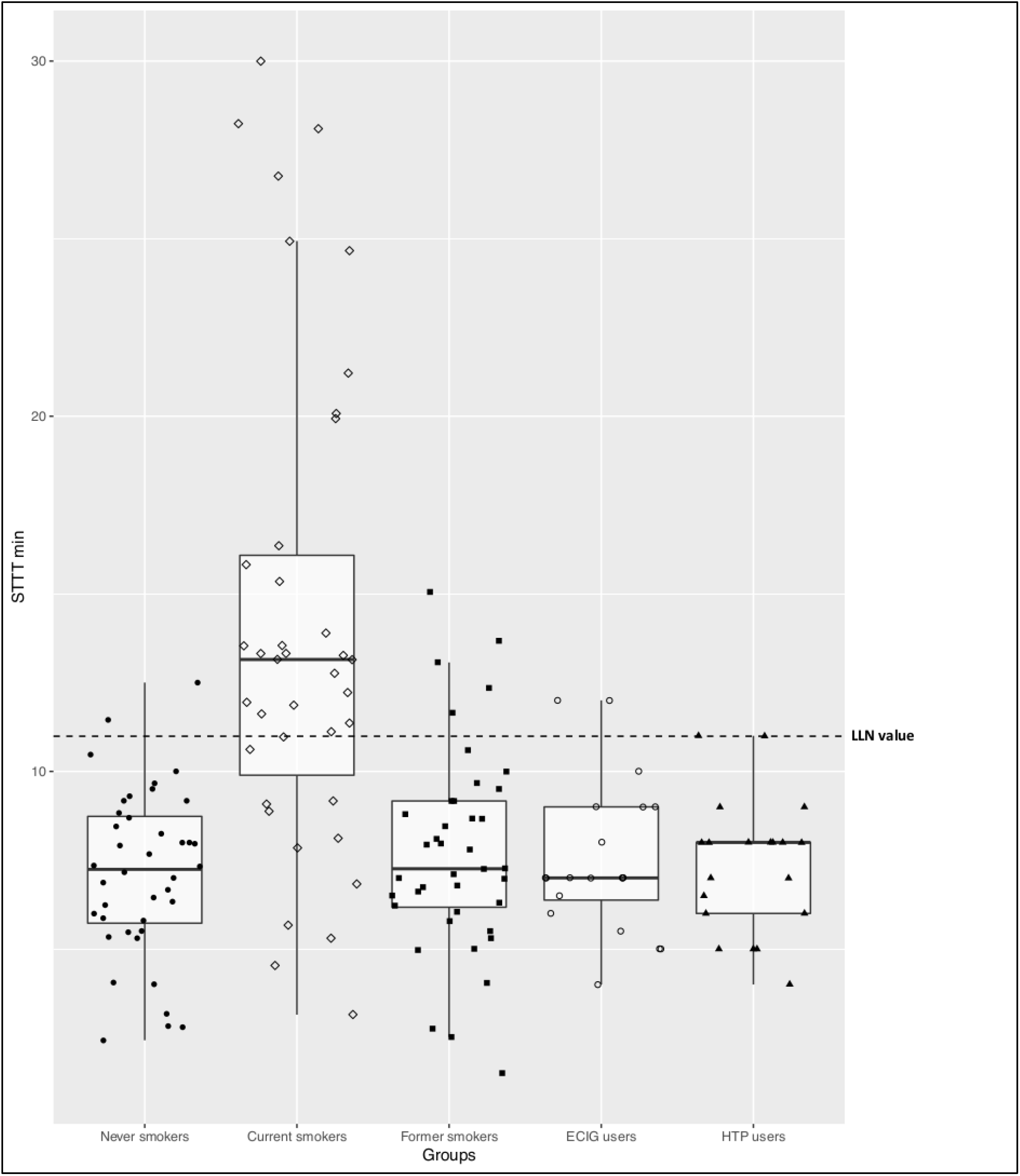
STTT measurements among never smokers (closed circles), current smokers (open diamonds), former smokers (closed squares), exclusive e-cigarette users (open circles), and exclusive heated tobacco product users (closed triangles). The median STTT (IQR) was prolonged only in Current Smokers, compared to other study groups. The calculated Lower Limit of Normality (LLN) was 10.99 minutes. The overall P value was calculated by Kruskal-Wallis test.

Pairwise comparisons between each study group showed that significant differences occurred only when current smokers were compared to any other study group, whereas no significant differences were observed for any other between-group comparisons (**Table 2**).

**Table 2.**
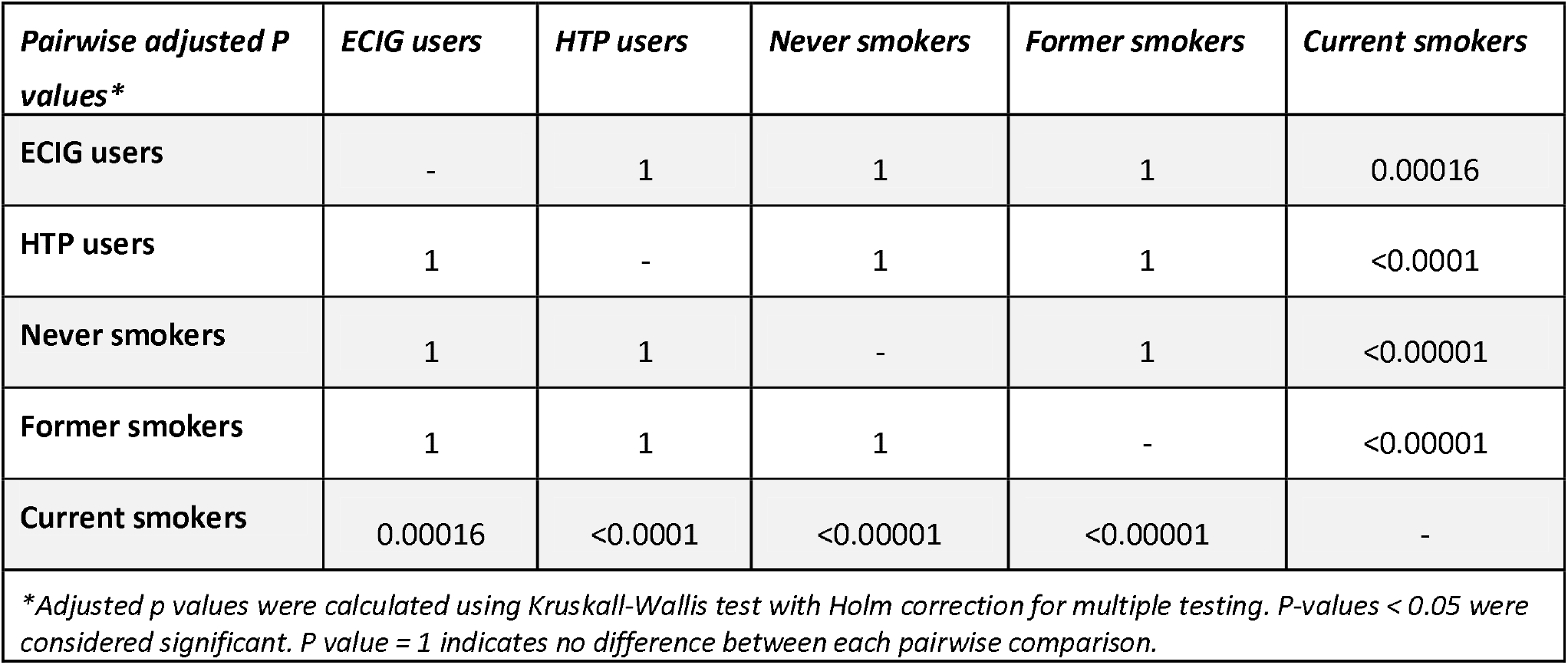
Pairwise Comparisons between Study Groups

The calculated Lower Limit of Normality (LLN) of 10.99 min in never smokers was used as a cut-off point for abnormal MCCTT measurements. As expected, most of current smokers (27/39; 69.2%) had an MCCTT value above the LLN, whereas only 12.5% (5/40) former smokers, 10% (2/20) exclusive EC users, and 10% (2/20) exclusive HTP users had MCCTT values above the LLN.

## DISCUSSION

This is the first study to investigate saccharin transit time in e-cigarettes and heated tobacco products users. Compared to never and former smokers, saccharin transit time in current cigarette smokers was nearly twice as long (7.24 and 7.26 min vs 13.15 min) and remarkably similar transit times were also observed in ECs (7.00 min) and HTPs users (8.00 min). Moreover, 90% of transit time measurements taken from EC users or HTP users were well within the lower limit of normal.

With a saccharin transit time that is similar to that of former and never smokers, EC and HTP users exhibit no significant impairment of MCC. Considering that exclusive EC and HTP users in our study have been only recently abstaining from smoking (3 to 36 months), complete MCC restoration after smoking cessation occurs quickly after quitting. Prospective studies are required to clarify the time-course of MCCTT restoration after smoking cessation.

In agreement with the findings of this paper, cigarette smoking slows down MCC and abstaining from smoking quickly restores MCC efficiency (16,17). However, that ECs and HTPs regular users exhibited no lengthening of saccharin test transit time indicates that combustion-free nicotine delivery technologies are unlikely to have detrimental effects on MCC function. This is a novel finding and requires explanation.

Whereas chronic exposure to toxic chemicals generated during tobacco combustion is known to cause functional alterations and structural damage of ciliated airway epithelial cells (3-6), very little is known about the effect of aerosol emissions from combustion-free nicotine delivery technologies. By completely substituting ECs for combustible tobacco cigarettes, users’ exposure to numerous toxicants and carcinogens present in combustible tobacco cigarettes is greatly reduced (19-21). Same marked reductions in toxicants exposures have been reported for HTPs compared to cigarettes (22,26,27).

Primary human bronchial epithelial cells exposed to cigarette smoke showed a clear reduction in mucus-secreting cells and their secretion activity as well as in cilia beating, with much less pronounced effects for the cells treated with EC aerosol (28). In an experimental model of excised bullfrogs’ palates, although exposure to EC aerosol emission had a modest inhibitory effect on mucus transport velocity, tobacco smoke exposure of the palates had a remarkable inhibitory effect (29). In a whole-body intense exposure protocol, no significant slowing in MCC by radioisotope technique was observed in mice exposed for 1 week to high levels of EC emission aerosols and trachea histology of sacrificed animals showed no apparent damage of the ciliated epithelial cells (30). Taken together these observations may suggest that combustion-free nicotine delivery technologies are unlikely to have detrimental effects on MCC function and add to the evidence that these products do not appear to pose a significant respiratory health hazard (31).

Some of the strengths of this study included: 1) exclusion of participants with any condition that could interfere with the results of saccharin test; 2) careful characterization of participants via detailed smoking, vaping, and HTP use history; 3) biochemical verification of participants’ smoking status by exhaled CO; 4) meticulous preparation and competent conduct of our standardized saccharin test, and 5) being one of the largest MCCTT study ever conducted.

When interpreting the study findings, many factors need to be considered. First, the reported lack of difference when comparing small study groups (i.e. EC and HTP) should be interpreted with caution. Yet, power analysis of the collected data indicates that a sample of at least 20 subjects for each group is adequately powered to detect significant differences in MCCTT. Moreover, careful examination of the individual saccharin test data on a case-by-case basis revealed identical 90% distribution of the measurements within the LLN value for both EC and HTP. Remarkably, significant differences were always reported when current smokers were included in all pairwise comparisons confirming the superior discriminatory capability of the saccharin test. Second, EC and HTP users in this study had relatively short duration of exposure (vaping and HTP usage history ranging from 3 to 36 months) that may have not been sufficient to show an effect. In addition, no EC user consumed more than 5 ml e-liquid/day and no HTP users puffed more than 15 sticks/day. Duration (years) and intensity of smoking (cigarettes/day) are significant predictors of MCCTT impairment among smokers, with high intensity smoking (heavy smokers: average of 39 cigs/day) nearly doubling MCCTT, and low smoking intensity (light smokers: average of 9 cigs/day) not having much of an effect (32). MCC impairment is also dependent on smoking duration, with smoking >5 years showing twice the saccharin transit time compared to smokers with a much shorter smoking history and to non-smokers (33). Thus, it is possible that the reported lack of impact on saccharin transit time among EC and HTP user could have been due to low-intensity aerosol exposure and consequently the possibility that longer exposure time and heavier consumption behaviours may cause significant MCC impairment should not be overlooked. Larger studies of inveterate users are required. Thirds, another limitation is that study samples consisted of relatively young subjects and their response to the saccharin test may not be representative of the general population. Consequently, firm conclusions cannot be drawn from the results and additional studies with more representative age groups are needed. Fourth, with the exception of a single GLO user, all other participants in the HTP study group were using IQOS. Therefore, HTP results are essentially product specific and cannot be extended to other HTP products. Conversely, EC users were consuming different types of vaping products. Fifth, although the results about ECs and HTPs appear to support tobacco industry data and may be therefore considered with reluctance, it is important that the data reported by the industry is independently verified by academic research.

In relation to the wider implications of this study, it is our opinion that measurement of saccharin transit time can be used as a sensitive biomarker of physiological effect for the detection of early respiratory health changes in smoking cessation studies and switching trials and to substantiate risk reduction and harm reversal of present and future combustion-free nicotine/tobacco products.

## Data Availability

The data that support the findings of this study are available from the corresponding author, RP, upon reasonable request.

## REFERENCES

1. US Department of Health and Human Services. The health consequences of smoking: 50 years of progress: a report of the surgeon general. Atlanta: US Department of Health and Human Services, Centers for Disease Control and Prevention, National Center for Chronic Disease Prevention and Health Promotion, Office on Smoking and Health. 2014.

2. U.S. Department of Health and Human Services. How tobacco smoke causes disease: the biology and behavioral basis for smoking-attrib-utable disease: a report of the surgeon general. Atlanta: U.S. Department of Health and Human Services, Centers for Disease Control and Prevention, National Center for Chronic Disease Prevention and Health Promotion, Office on Smoking and Health. 2010.

3. Sisson JH, Papi A, Beckmann JD, Leise KL, Wisecarver J, Brodersen BW, Kelling CL, Spurzem JR, Rennard SI. Smoke and viral infection cause cilia loss detectable by bronchoalveolar lavage cytology and dynein ELISA. Am J Respir Crit Care Med 1994;149:205–213.

4. Jacob S, Kraft R, Zhu Y, Jacob RK, Herndon DN, Traber DL, Hawkins HK, Cox RA. Acute secretory cell toxicity and epithelial exfoliation after smoke inhalation injury in sheep: an electron and light microscopic study. Toxicol Mech Methods 2010 Oct;20(8):504–9.

5. Simet SM, Sisson JH, Pavlik JA, Devasure JM, Boyer C, Liu X, Kawasaki S, Sharp JG, Rennard SI, Wyatt TA. Long-term cigarette smoke exposure in a mouse model of ciliated epithelial cell function. m J Respir Cell Mol Biol. 2010 Dec; 43(6):635–40.

6. Kuehn D, Majeed S, Guedj E, Dulize R, Baumer K, Iskandar A, Boue S, Martin F, Kostadinova R, Mathis C, Ivanov N, Frentzel S, Hoeng J and Peitsch MC (2015) Impact assessment of repeated exposure of organotypic 3D bronchial and nasal tissue culture models to whole cigarette smoke. Journal of Visualized Experiments, 96:e52325. doi:10.3791/52325.

7. Cohen D, Arai SF, Brain JD. Smoking impairs long-term dust clearance from the lung. Science 1979;204:514–517.

8. Domagala-Kulawik J. Effects of cigarette smoke on the lung and systemic immunity. J Physiol Pharmacol. 2008 Dec; 59 Suppl 6():19–34.

9. Rodgman A, Perfetti TA. The chemical components of tobacco and tobacco smoke. Boca Raton (FL): CRC Press, Taylor & Francis Group; 2009.

10. Dalhamn T, Rylander R. Ciliotoxicity of cigar and cigarette smoke. Arch Environ Health. 1970;20:252–253.

11. Cosio MG, Hale KA, Niewoehner DE. Morphologic and morphometric effects of prolonged cigarette smoking on the small airways. Am Rev Respir Dis. 1980;122(2):265–21.

12. Konrad F, Schreiber T, Brecht-Kraus D, Georgieff M. Mucociliary transport in ICU patients. Chest. 1994;105(1):237–41.

13. Prescott E, Lange P, Vestbo J. Chronic mucus hypersecretion in COPD and death from pulmonary infection. Eur Respir J. 1995;8(8):1333–8.

14. Lawrence H, Hunter A, Murray R, Lim WS, McKeever T. Cigarette smoking and the occurrence of influenza - Systematic review. J Infect. 2019;79(5):401–406. doi:10.1016/j.jinf.2019.08.014

15. Pagliuca G, Rosato C, Martellucci S, de Vincentiis M, Greco A, Fusconi M, et al. Cytologic and functional alterations of nasal mucosa in smokers: temporary or permanent damage? Otolaryngol Head Neck Surg. 2015; 152(4):740–5.

16. Ramos EM, De Toledo AC, Xavier RF, et al. Reversibility of impaired nasal mucociliary clearance in smokers following a smoking cessation programme. Respirology. 2011;16(5):849–855.

17. Utiyama DM, Yoshida CT, Goto DM, et al. The effects of smoking and smoking cessation on nasal mucociliary clearance, mucus properties and inflammation. Clinics (Sao Paulo). 2016;71(6):344–350.

18. Polosa R, Farsalinos K, Prisco D. Health impact of electronic cigarettes and heated tobacco systems. Intern Emerg Med. 2019;14(6):817–820.

19. Farsalinos KE and Polosa R. Safety evaluation and risk assessment of electronic cigarettes as tobacco cigarette substitutes: a systematic review. Ther Adv Drug Saf 2014; 5: 67–86.

20. Daynard, R. (2018). Public health consequences of e-cigarettes: a consensus study report of the National Academies of Sciences, Engineering, and Medicine. Journal of Public Health Policy. https://doi.org/10.1057/s41271-018-0132-1

21. McNeill, A., Brose, L.S., Calder, R., Bauld. L. and Robson, D. (2018), Evidence Review of E-Cigarettes and Heated Tobacco Products, 2018. A Report Commissioned by Public Health England, Public Health England, London. <https://www.gov.uk>/government/publications/e-cigarettes-and-heated-tobacco-products-evidence-review

22. Caponnetto, P., Maglia, M., Prosperini, G., Busà, B. and Polosa, R. (2018), “Carbon monoxide levels after inhalation from new generation heated tobacco products”, Respiratory Research, Vol. 19 No. 1, pp. 1–4.

23. O’Leary R, Polosa R. Tobacco Harm Reduction in the 21^st^ Century. Drugs and Alcohol Today 2020 (in press).

24. Sherly D, Prathibha KM (2014). Measurement of Nasal Mucociliary Clearance. Clin Res Pulmonol. ;2(2):1019.

25. https://www.>cdc.gov/nchs/nhis/tobacco/tobacco_glossary.htm

26. Mallock N, Böss L, Burk R, et al. Levels of selected analytes in the emissions of “heat not burn” tobacco products that are relevant to assess human health risks. Arch Toxicol. 2018;92(6):2145–2149. doi:10.1007/s00204-018-2215-y

27. Iskandar AR, Zanetti F, Kondylis A, et al. A lower impact of an acute exposure to electronic cigarette aerosols than to cigarette smoke in human organotypic buccal and small airway cultures was demonstrated using systems toxicology assessment. Intern Emerg Med. 2019;14(6):863–883. doi:10.1007/s11739-019-02055-x

28. Aufderheide M, Emura M. Phenotypical changes in a differentiating immortalized bronchial epithelial cell line after exposure to mainstream cigarette smoke and e-cigarette vapor. Exp Toxicol Pathol. 2017;69(6):393–401.

29. Palazzolo DL, Nelson JM, Ely EA, Crow AP, Distin J, Kunigelis SC. The Effects of Electronic Cigarette (ECIG)-Generated Aerosol and Conventional Cigarette Smoke on the Mucociliary Transport Velocity (MTV) Using the Bullfrog (R. catesbiana) Palate Paradigm. Front Physiol. 2017;8:p1023. Published 2017 Dec 11. doi:10.3389/fphys.2017.01023

30. Laube BL, Afshar-Mohajer N, Koehler K, et al. Acute and chronic in vivo effects of exposure to nicotine and propylene glycol from an E-cigarette on mucociliary clearance in a murine model. Inhal Toxicol. 2017;29(5):197–205. doi:10.1080/08958378.2017.1336585

31. Polosa R, O’Leary R, Tashkin D, Emma R, Caruso M. The effect of e-cigarette aerosol emissions on respiratory health: a narrative review. Expert Rev Respir Med. 2019; 13(9):899–915. doi:10.1080/17476348.2019.1649146

32. Xavier RF, Ramos D, Ito JT, et al. Effects of cigarette smoking intensity on the mucociliary clearance of active smokers. Respiration. 2013;86(6):479–485. doi:10.1159/000348398

33. Baby MK, Muthu PK, Johnson P, Kannan S. Effect of cigarette smoking on nasal mucociliary clearance: A comparative analysis using saccharin test. Lung India. 2014;31(1):39–42. doi:10.4103/0970-2113.125894

